# Self-assessment of COVID-19 vaccination efficacy using a lateral flow tests for SARS-CoV-2 S1 protein antibody

**DOI:** 10.1101/2021.06.27.21258591

**Authors:** Maohua Li, Yi Shan, Kun Cai, Wenlin Ren, Hunter Sun, Shujiang Wu, Jianli Li, Dee Hong, Zhenxing Zhang, Qi Wang, Lijun Qin, Yufei Sun, Chunsheng Ye, Huan Jiang, Zhenyu Wang, Yongzhong Jiang, Chao Liu, Bin Hu, Ruifeng Chen, Le Sun

## Abstract

**Background:** More than ten novel COVID-19 vaccines have been approved with protections against SARS-CoV-2 infections ranges between 52-95%. It is of great interest to the vaccinees who have received the COVID-19 vaccines, vaccine developers and authorities to identify the non-responders in a timely manner so intervention can take place by either giving additional boosts of the same vaccine or switching to a different vaccine to improve the protection against the SARS-CoV-2 infections. A robust correlation was seen between binding antibody titer and efficacy (p=0.93) in the clinic studies of 7 COVID-19 vaccines, so it is of urgency to develop a simple POCT for vaccinees to self-assess their immune response at home.

**Methods:** Using CHO cell-expressed full length SARS-CoV2 S1 protein as coating antigen on colloidal gold particles, a SARS-CoV-2 S1 IgG-IgM antibody lateral flow test kit (POCT) was developed. The test was validated with negative human sera collected prior to the COVID-19 outbreaks, and blood samples from human subjects prior, during, and post-immunization of COVID-19 vaccines.

**Results:** The specificity of the POCT was 99.0%, as examined against 947 normal human sera and 20 whole blood samples collected pre-immunization. The limit of detection was 50 IU/mL of pseudovirus neutralizing titer (PVNT) using human anti-SARS-2 neutralizing standards from convalescent sera. The sensitivity of POCT for SARS-CoV-2 S1 protein antibody IgG-IgM was compared with SARS-CoV-2 RBD antibody ELISA and determined to be 100% using 23 blood samples from vaccinated human subjects and 10 samples from non-vaccinated ones. Whole blood samples were collected from 119 human subjects (ages between 22-61 years) prior to, during, and post-vaccination of five different COVID-19 vaccines. Among them, 115 people tested positive for SARS-CoV-2 S1 antibodies (showing positive at least once) and 4 people tested negative (tested negative at least twice on different days), demonstrating 96.64% of seroconversion after full-vaccination. 92.3% (36/39) of the human subjects who were younger than 45 achieved seroconversion within 2 weeks while only 57.1% (4/7) of subjects older than 45 tested positive for S1 antibodies, suggesting that younger people develop protection much faster than older ones. Even though the S1 antibody level in 88% of human subjects vaccinated with inactivated virus dropped below 50 IU/mL two months later, one boost could quickly raise the S1 antibody titer above 50 IU/mL of PVNT, indicates that the initial vaccination was successful and immunization memory was developed.

**Conclusion:** Using the lateral flow tests of SARS-CoV2 S1 IgG+IgM, vaccinated human subjects can easily self-assess the efficacy of their vaccination at home. The vaccine developer could quickly identify those non-responders and give them an additional boost to improve the efficacy of their vaccines. Vaccinees who failed in response could switch to different types of COVID-19 vaccines since there are more than 10 COVID-19 vaccines approved using three different platform technologies.

**Highlights:**

1. More than ten novel COVID-19 vaccines have been approved with protections against SARS-CoV-2 infections ranges between 52-95%. It is of great interest to the vaccinees who have received the COVID-19 vaccines, vaccine developers and authorities to identify the non-responders in a timely manner.
2. A highly specific and very simple lateral flow test kit for measurement of SARS-CoV-2 S1IgG+IgM antibodies post-immunization of COVID-19 vaccine using peripheral blood was developed as a home-test assay with a limit of detection (LOD) at 50 IU/mL of pseudovirus neutralizing titer (PVNT).
3. After full vaccinations with COVID-19 vaccines, 96.6% of the volunteers successfully achieved the seroconversion of SARS-CoV-2 S1 IgG+IgM antibody.
4. 92.3% (36/39) of the human subjects who were younger than 45 achieved seroconversion within 2 weeks while only 57.1% (4/7) of subjects older than 45 tested positive for S1 antibodies, suggesting that younger people develop protection much faster than older ones.
5. Even though the S1 antibody level in 88% of human subjects vaccinated with inactivated virus dropped below the detection 2-6 months later, one boost could quickly raise the S1 antibody titer above 50 IU/mL of PVNT, indicating that the initial vaccination was successful and immunization memory was developed.

## Introduction

As of May 10^th^, there were 159,117,886 confirmed cases of COVID-19 with 3,303,986 deaths in the world and the numbers of infections were growing at a pace of around 600,000 case per day^1^. Though more than 10 novel COVID vaccines have been approved under emergency use with demonstrated efficacy against the wild-type SARS-CoV-2 virus, additional affordable and deliverable vaccines are needed to meet the unprecedented global needs. It is also of great urgency to develop New COVID-19 vaccines targeting new variants of concern. At the same time, with placebo-controlled efficacy trials becoming infeasible due to the introduction of approved vaccines, a correlate of protection (CoP) is urgently needed to provide a path for regulatory approval of new COVID-19 vaccines. After a thorough analysis of data from 7 approved COVID-19 vaccines, Dr. K. Earle and her co-workers found that there was a better correlation between S/RBD binding antibody ELISA titer and efficacy (p=0.93) than neutralizing titer and efficacy (p=0.79) ^2^. Their report supports the use of post-immunization binding antibody levels as the basis for a CoP.

WHO recently authorized the emergency use license (EUL) of Sinopharm’s BBIBP COVID-19 vaccine at the end of April 2021^3^. The overall efficacy of BBIBP COVID-19 vaccine in protection against infection was around 78%^4^. At this moment, it is unclear why the 22% vaccinees were found infected by SARS-CoV-2 after full vaccination. Possibilities include in-effectiveness of vaccination or the quick decay of immune response.

Previously we have developed a highly specific and very sensitive serological SARS-CoV-2 antibody assay with an overall accuracy of 97.3% using CHO-expressed SARS-CoV-2 S1 protein for screening of SARS-CoV-2 infection^5^. The assay was able to detect SARS-CoV2 S1 antibody on day one after the onset of COVID-19 disease. SARS-CoV-2 S1 antibodies were detected in 28 out of 276 asymptomatic medical staff and 1 out of 5 nucleic acid test-negative “Close contacts” of a COVID-19 patient^5^. However, the assay has to be performed by skilled technicians in the laboratory and is not suitable for monitoring immune responses in vaccinees in a timely manner.

In this paper, a lateral flow test of SARS-CoV-2 S1 IgG+IgM was developed. It offers a tool to vaccinees for self-assessment of immune responses and persistence of COVID-19 vaccination, reducing the risk of infections due to ineffective vaccinations and/or quick decay of SARS-CoV-2 antibodies. It could also help to quickly identify those human subjects who failed to develop enough immune response after the full vaccination, allowing an additional boost or switch to another type of COVID-19 vaccine.

## Materials and methods

### Regents and supplies

SARS-CoV-2 neutralizing antibody standard (SB6N83NP) was from National Inst. for Food and Drug Control, Beijing, China. SARS-CoV-2 S1-His6X and RBD-Dimer-6XHis recombinant proteins were made by ZhenGe Biotech., Shanghai, China. High-binding 96-well ELISA plates were purchased from Corning, USA. Goat anti-human IgG (H+L) peroxidase conjugate was sourced from Jackson Immunoresearch, USA.

### Blood samples for assay

Strong human negative plasma samples were collected prior to the COVID-19 outbreak. Human negative whole blood samples were collected from either non-vaccinated persons or just prior to the vaccination of COVID-19 vaccines. 10 µL of blood was drawn from fingertips by the volunteers themselves. Human subjects were immunized with one of five different COVID-19 vaccines including inactivated virus vaccines (CoronaVac from Sinovac, BBIBP-CorV from SinoPharm. and one from Inst. Med. Biol. Chinese Acad. of Med. Sci.), Adenovirus vaccine (Ad5-nCoV from Cansino) and recombinant protein vaccine (ZF2001 from Zhifei Biol.). Informed consent was obtained from all the human subjects who participated in the study and the protocols were approved by the institutional ethical committee.

### Rapid Immunochromatographic Assay

Mouse anti-human IgG antibody (BCAB-M ANTIHIgG-AW, 1.5mg/mL) was dispensed on a nitrocellulose membrane as a “IgG test” (IgG) line with a speed of 0.8 µL/cm, mouse anti-human IgM antibody (BCAB-M ANTIHIgM-AW, 0.8mg/ml) was dispensed on a nitrocellulose membrane as a “IgM test” (IgM) line with a speed of 0.8 µL/cm, and 30% of BSA conjugated with latex particles and 2mg/ml of green pigment were fixed as a “control” (C) line. The sprayed nitrocellulose membrane was dried for more than 12 hours at 45°C and stored for future use. Colloidal gold particles were conjugated with the SARS-CoV-2 Spike 1 protein to prepare antigen conjugates, and conjugate solution was sprayed evenly on a glass fiber at 2 µL/cm. This was followed by drying for more than 12 hours at 37°C. Finally, the sample pad, conjugate pad, nitrocellulose membrane, and absorbent pad were attached to a lamination pad with pressure-sensitive adhesive, and then cut to a width of 4 mm to install in a plastic cassette.

### SARS-CoV-2 RBD Antibody ELISA kit

Briefly, recombinant SARS-CoV-2 RBD-Dimer-6XHis was diluted in PBS (10 mM, pH 7.4) and 100µL of the solution was added to each well of 96-well ELISA plates and incubated overnight at 2-8°C. The wells were emptied and washed twice with PBS and unsaturated sites were blocked with 3% BSA in PBS by incubating for 1 hour at room temperature. Coated plates were air-dried and sealed in plastic bags and stored at 2-8°C until use.

For ELISA, each serum sample was tested in duplicate. Prior to testing, human samples or the standards were diluted 1:15 in sample dilution buffer (20% Calf serum in PBS). 100µL of appropriately diluted sample was added to each well of the RBD-Dimer-6XHis-coated plates and incubated for 0.5 hour at 37°C with constant shaking. The wells were emptied and washed twice with washing buffer. 100 µL of appropriately diluted HRP-conjugated goat anti-human IgG (H+L) was added to each well and incubated for another 15 min at 37°C with constant shaking. The wells were emptied and washed five times before addition of TMB substrate solutions. The chromogenic development was stopped using 0.1M H_2_SO_4_ after 5-10 minutes of incubation in the dark. Optical density (OD) was measured at 450nm wave length in a microplate spectrophotometer (Thermo Scientific, Multiskan MK3).

## Results

### Validation of POCT for SARS-CoV-2 S1 protein antibody IgG-IgM

The diagnostic specificity of the POCT was demonstrated by testing 967 human samples including 947 human serum samples collected prior to the outbreak of COVID-19 (strong negatives) and 20 whole blood samples collected prior to immunization with COVID-19 vaccines (negatives). As shown in Table 1-1, while 10 of the 947 strong negatives showed false-positives, none of the 20 negative samples collected prior to immunization tested positive. Therefore, the overall specificity of the POCT for SARS-CoV-2 S1 protein antibody IgG-IgM was 99.0%.

**Table 1.**
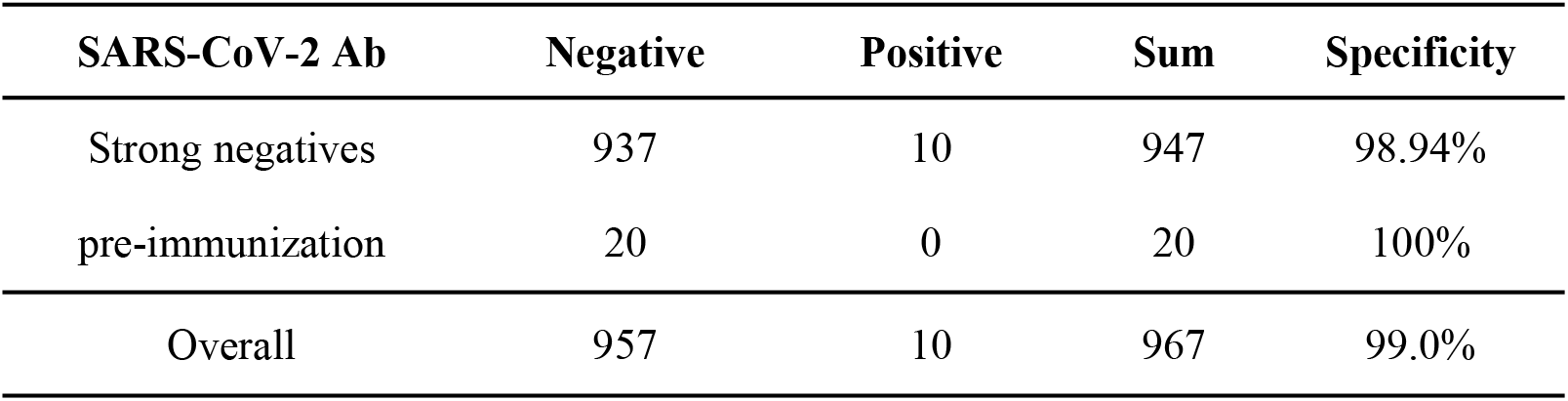
Specificity of the POCT for SARS-CoV-2 S1 protein antibody IgG-IgM Strong human negative plasma samples were collected prior to the COVID-19 outbreak. Human negative whole blood samples were collected from either non-vaccinated persons or just prior to the vaccination of COVID-19 vaccines. SARS-CoV-2 Ab-Negative indicates no SARS-CoV-2 antibody detected in the sample. SARS-CoV-2 Ab-Positive indicates SARS-CoV-2 antibody was detected in the sample.

| SARS-CoV-2 Ab | Negative | Positive | Sum | Specificity |
| --- | --- | --- | --- | --- |
| Strong negatives | 937 | 10 | 947 | 98.94% |
| pre-immunization | 20 | 0 | 20 | 100% |
| Overall | 957 | 10 | 967 | 99.0% |

The limit of detection of the POCT was examined using a human anti-SARS-2 neutralizing antibody standard purchased from the National Inst. for Food and Drug Control. The neutralizing antibody standard was pooled-sera from convalescent COVID-19 patients with a pseudovirus neutralizing titer (PVNT) of 1000 IU/mL. The neutralizing antibody standards were diluted with normal human sera to various concentrations between 1-1000 IU/mL. As shown in Fig. 1, the LOD of the POCT was determined at 50 IU/mL of PVNT. Surprisingly, a strong IgM band of S1 antibodies was detected in the pooled sera from the convalescent COVID-19 patients, suggesting either some of the convalescent patients still carried the virus or the half-life of COVID-19 IgM is much longer than we expected.

**Fig. 1.**
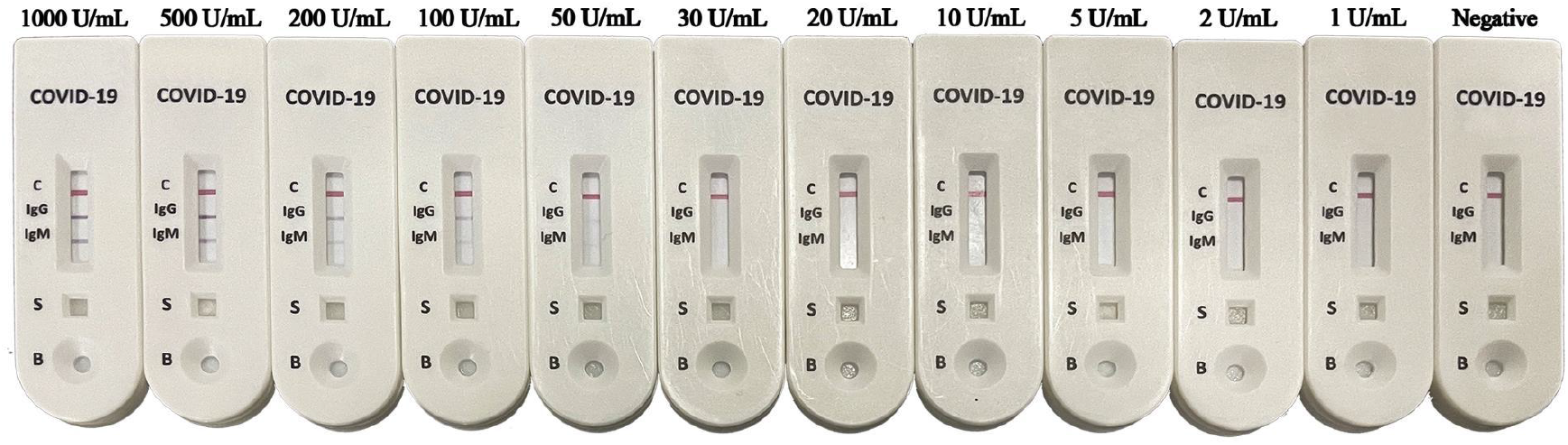
LOD of the POCT for SARS-CoV-2 S1 protein antibody IgG-IgM. A neutralizing antibody standard from the National Inst. for Food and Drug Control was diluted with human normal sera to various concentrations between 1-1,000 IU/mL. 5 µL each of the diluted standards was applied to the sample loading well (S) first, and two drops of the buffer were added to the buffer well (B). After 5-20 mins, photographs were taken.

The sensitivity of POCT for SARS-CoV-2 S1 protein antibody IgG-IgM was compared with SARS-CoV-2 RBD antibody ELISA. A total of 33 blood samples were collected from COVID-19-vaccinated human subjects prior and post full vaccination. As shown in Table 2, out of the 23 samples tested positive for S1 protein antibody using the POCT, 21 were RBD antibody ELISA-positive and two were equivocal. None of the POCT negatives was RBD antibody ELISA-positive. These data suggest the sensitivity of POCT is close to 100%. The slight difference observed between the two assays may be due to the different antigens used.

**Table 2.** Sensitivity of POCT for SARS-CoV-2 S1 protein antibody IgG-IgM Samples were collected from human subjects prior to and after full vaccination of COVID-19 vaccines. The samples were tested side-by-side with SARS-CoV-2 RBD Antibody ELISA and SARS-CoV-2 S1 IgG+IgM POCT. For POCT, band(s) of IgG and/or IgM visible within 20 mins were defined as positive. For ELISA, if S/N≥2.0 was defined as RBD positive, between 1.5-1.9 was defined as RBD Ab Equivocal Positive, and <1.5 was considered Negative.

|  | **POCT**<br>**Positive** | **POCT**<br>**Negative** | **Sum** |
| --- | --- | --- | --- |
| **RBD Ab Positive** | 21 | 0 | 21 |
| **RBD Ab Equivocal Positive** | 2 | 0 | 2 |
| **RBD Ab Negative** | 0 | 10 | 10 |
| **Sum** | 23 | 10 | 33 |

### Self-examination of SARS-CoV-2 S1 antibody IgG-IgM level post vaccination

By the end of April of 2021, NMPA had conditionally approved 5 COVID-19 vaccines, including 3 inactivated virus vaccines, one adenovirus carrying SARS-CoV-2 S protein gene and one using recombinant SARS-CoV-2 RBD-Dimer. By now, more than 300 million doses of COVID-19 vaccines have been administered. To monitor the immunogenicity of the approved COVID-19 vaccines, more than 2,000 SARS-CoV-2 S1 antibody IgG-IgM POCT kits were sent to people in more than 50 cities and counties in mainland China. Whole blood samples were self-collected from fingertips by the volunteers prior to, during, and post-vaccination of five different approved COVID-19 vaccines. Close to 300 testing results with pictures were received and majority of them were from human subjects between the ages of 18 and 59 in the working force.

To determine the best blood sampling time, for the first few human subjects, multiple blood drawings were done during and after the full vaccination. When we looked into the time to seroconversion, exampled in Fig. 2, none of the blood samples drawn from human subjects found SARS-CoV-2 S1 IgG+IgM positive prior to full vaccination of SinoPharm’s CorV COVID-19 vaccine.

**Fig. 2.**
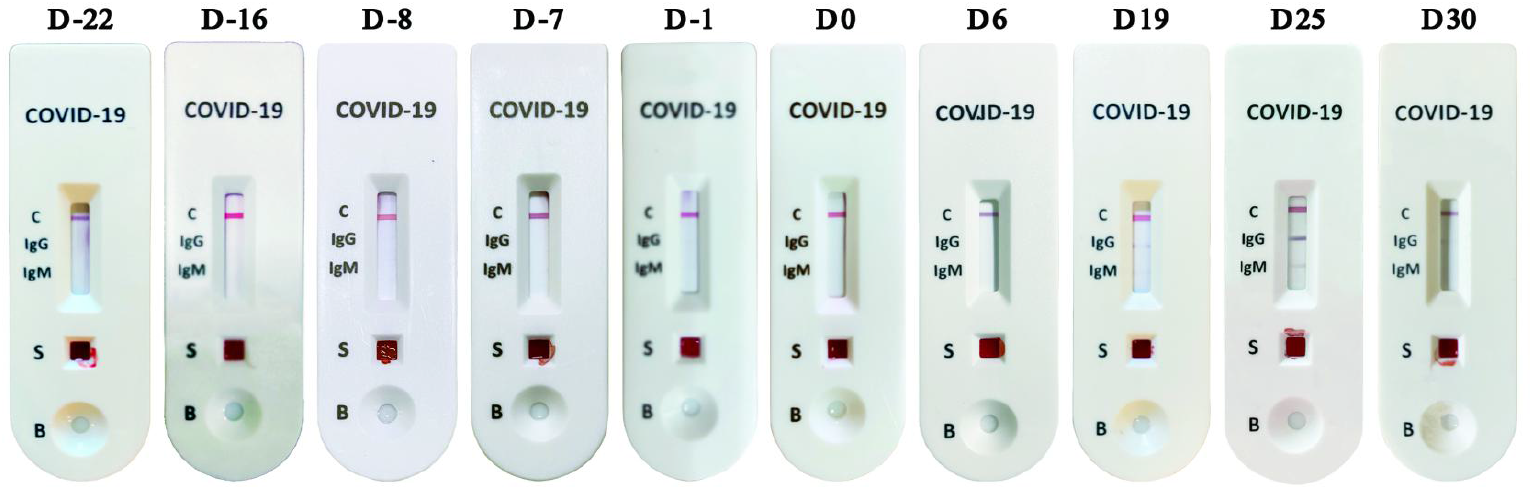
Time course of SARS-CoV-2 S1 antibody IgG-IgM. Samples were collected from one male human subject at different time points prior, during and post full vaccination of SinoPharm’s CorV COVID-19 vaccine. Band(s) of IgG and/or IgM visible within 20 mins was determined as positive.

Based on these data, to reduce subsequent unnecessary blood drawing, no sampling was taken prior to the completion of full vaccination of inactivated virus vaccine. For one-dose adenovirus-based vaccine, according to Manufacture’s suggestion, blood samples were collected 28 days post the immunization.

As shown in Table 3, for 119 subjects who completed the full vaccinations, 115 people tested positive for SARS-CoV-2 S1 antibodies (at least showed positive once) and 4 persons tested negative (tested negative at least twice at different days), demonstrating a 96.6% seroconversion rate. Our data is in strong agreement with what has been reported for seroconversion rates for SARS-CoV-2 pseudo virus neutralizing assays or SARS-CoV-2 S/RBD protein binding antibody ELISA data from Phase I and Phase II clinical trials for this age group^6,7,8^.

**Table 3.** Seroconversion rates for S1 antibodies Samples were collected from human subjects at least one week after full vaccination of COVID-19 vaccines. Band(s) of IgG and/or IgM visible within 20 mins was determined as positive.

|  | Fully<br>vaccination | S1 Ab<br>positive | S1 Ab<br>negative | Seroconversion |
| --- | --- | --- | --- | --- |
| Overall | 119 | 115 | 4 | 96.64% |
| <45 | 89 | 87 | 2 | 97.77% |
| ≥45 | 30 | 28 | 2 | 93.33% |

## Discussion

With close to 300 test results returned with photographs, we observed certain patterns, such as the negative impact of aging on the immune responses. As shown in Table 4-1, within 2 weeks (D1-14) post-full immunization, for age younger than 45 years old, 92.3% (36/39) showed S1 antibody seroconversion (≥50 IU/mL of PVNT). However, only 57% (4/7) of the people who were older than 45 tested positive for S1 antibodies. Even though the elder group did catch up later with seroconversion rate at 93.3% between day 15-59 post full vaccination, the average of S1 antibody level was much lower (data not shown).

**Table 4-1.**
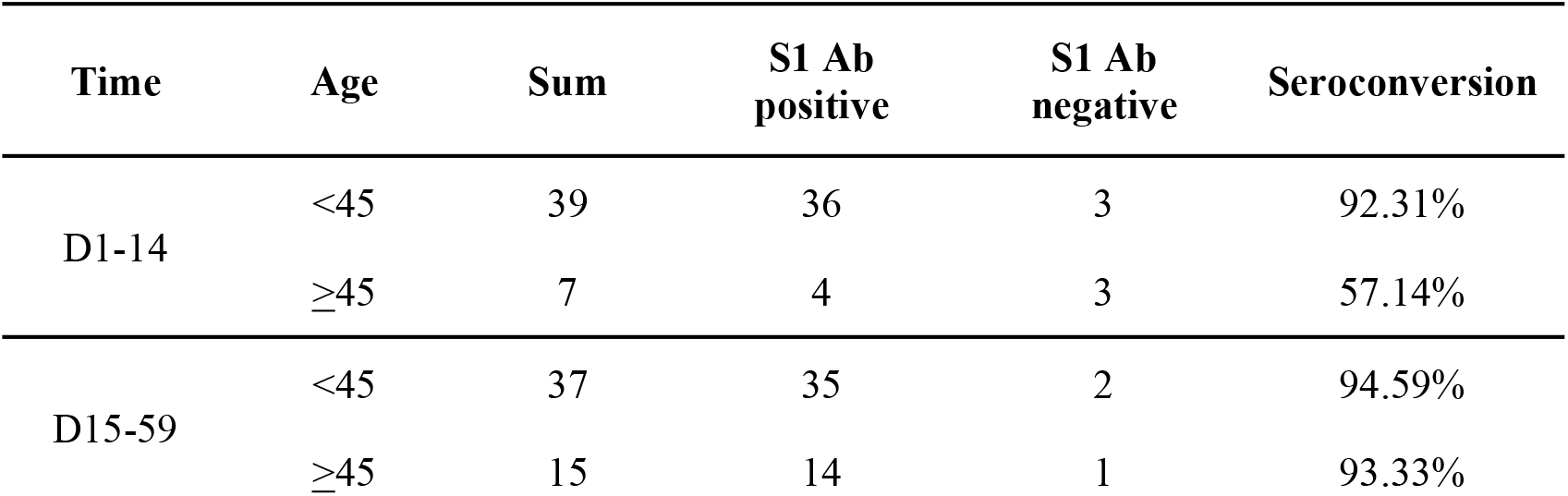
Time to seroconversion Samples were collected from human subjects at different time points post full vaccination. Band(s) of IgG and/or IgM visible within 20 mins was determined as positive.

| Time | Age | Sum | S1 Ab<br>positive | S1 Ab<br>negative | Seroconversion |
| --- | --- | --- | --- | --- | --- |
| D1-14 | <45 | 39 | 36 | 3 | 92.31% |
|  | ≥45 | 7 | 4 | 3 | 57.14% |
| D15-59 | <45 | 37 | 35 | 2 | 94.59% |
|  | ≥45 | 15 | 14 | 1 | 93.33% |

As shown in Table 4-2, for all 35 samples collected at least two months post the full vaccination, 42.8% (15/35) remained positive for S1 antibodies (above the 50 IU/mL). When we examine the data by ages, only 22% (2/9) of subjects 45 or older remained S1 antibody-positive two months later. In addition, we also noticed some of the elder vaccinees still had IgM of S1 antibodies 2 months or longer post-vaccination. This will actually create a problem to use SARS-CoV-2 IgM as a bio-marker to distinguish new infection from vaccination.

**Table 4-2.** Persistence of immune response in vaccinees Samples were collected from human subjects at least 2 months post-full vaccination. Band(s) of IgG and/or IgM visible within 20 mins were defined as positive.

| Type of vaccine | Age | Sum | S1 Ab positive | S1 Ab negative | Seroconversion |
| --- | --- | --- | --- | --- | --- |
| Overall | <45 | 26 | 13 | 13 | 50.0% |
|  | ≥45 | 9 | 2 | 7 | 22.2% |
| Inactivated virus | all | 25 | 3 | 22 | 12.00% |

As has been reported about the quick decay of S antibodies in convalescent COVID-19 patients^9^, in this study we found that only 12% (3/25) of the human subjects immunized with inactivated virus COVID-19 vaccine had S1 antibody above the 50 IU/mL level 2 months later post full immunization. Since the half-lives for different isotypes of human IgGs are very different, more studies are needed to examine the isotypes of S1 antibodies developed between different age groups after vaccination with inactivated virus COVID-19 vaccines.

Three of the vaccinees, whose S1 antibody titers dropped below detection of limit 3-month post-vaccination, received one boost and all of them showed S1 antibody IgG-positive within two weeks, suggesting that the past vaccination was successful and the memory of immunization was retained.

WHO recently authorized the emergency use license (EUL) of Sinopharm’s BBIBP COVID-19 and Sinovac’s CoronaVac. The overall efficacy of BBIBP COVID-19 vaccine in protection against infection was around 78%, while the reported protections of CoronaVac COVID-19 vaccine were between 50-70% in different countries. At this moment, it is unclear what were the causes of virus breakthrough. The regulatory agencies and vaccine developers were keen to know whether it is due to 1) ineffectiveness of vaccination, 2) the quick decay of immune response, or 3) immune escape of variants of concern.

Dr. JZ Wang’s group published a paper on EMI in March 2021 demonstrating that heterologous prime-boost could break the protective immune response bottleneck of COVID-19 vaccine in a mouse model^10^. A clinical trial (NCT04833101) with adenovirus-based Ad5-nCoV or inactivated virus vaccine CoronaVac as the priming vaccine and recombinant RBD-based ZF2001 as the boosting one was started in May 2021 in Jiangsu, China^11^.

With the POCT of SARS-CoV-2 S1 IgG+IgM we developed here, the vaccine developers can quickly identify the non-responders and give them an additional boost to improve the efficacy of their vaccines. To vaccinees who failed in immune response to their current vaccine, they could switch to different types of COVID-19 vaccines, since there are more than 10 COVID-19 vaccines approved using three different platform technologies including recombinant protein-based ones from Zhifei and Novavax, mRNA-based ones from Pfizer/BioNTech and Moderna, and Adenovirus-based ones from AZ/Oxford and Casino.

## Data Availability

The data used to support the findings of this study are available from the corresponding author.

## Author contributions

MHL, JLL, CL, QW contributed the development of RBD antibody ELISA and perform the assays. SJW, ZXZ, DH, YFS, CSY, HJ, XXW contributed the development of POCT for SARS-CoV-2 S1 IgG+IgM. RFC, WLR, YS, YZJ, KC, BH, LJQ contributed the blood sample data collecting and analysis. HS contributed market studies and technical writing.

## Acknowledgements

This work is supported by Research Grants from Beijing Science and Technology Commission, Bill & Melinda Gates Foundation, Zhejiang Quzhou Green Innovation Park, Tianjin Taida Economic Development Zone to Le Sun. We thank Dr. Lidong Guan from National Inst. for Food and Drug Control, China for providing SARS-CoV-2 neutralizing antibody standard (SB6N83NP). We thank Dr. David Barnes for English editing.

